# Real-World Serum Neurofilament Light Chain and GFAP in Amyotrophic Lateral Sclerosis on a Commercial ECLIA Platform

**DOI:** 10.64898/2026.06.24.26356370

**Authors:** Nicholas S. Streicher

## Abstract

**Background:** Neurofilament light chain (NfL) is an established prognostic biomarker in amyotrophic lateral sclerosis (ALS): SIMOA-based studies show that baseline serum NfL predicts ALSFRS-R slope and survival and serves as an outcome measure in clinical trials. The commercial Roche Elecsys electrochemiluminescence immunoassay (ECLIA) reads 6-to 8-fold lower than SIMOA, and its clinical utility in ALS is uncharacterized. We assessed whether ECLIA NfL retains this correlation in routine care and whether GFAP or S-100B helps.

**Methods:** Retrospective analysis of 58 chart-confirmed ALS patients at Georgetown University Hospital (2022–2026), biomarkers on the LabCorp Roche Elecsys ECLIA. The NfL–ALSFRS-R correlation was assessed where both measures fell within matching windows; serial NfL, in patients with repeat draws.

**Results:** First-per-patient NfL median was 7.06 pg/mL (IQR 4.06–17.30; CV 99%). Among 31 patients with matched NfL and ALSFRS-R decline rates, Spearman r = 0.704; within 90 days (n = 17), r = 0.809 (both p < 0.0001). Fast progressors (n = 8) had mean NfL 17.10 pg/mL versus 4.64 in slow progressors (n = 21), a 3.7-fold separation. Serial NfL captured rising trajectories and stable low values. GFAP scaled with age, not the disease, and did not significantly track progression rate, stage, or motor-neuron predominance; S-100B, assessed preliminarily, added little.

**Conclusions:** Commercial ECLIA brings NfL into routine ALS care; its prognostic correlation with progression rate survives real-world fragmentation. The actionable unit is the longitudinal trajectory, not the single value, read against platform-specific reference ranges and clinical context (genotype, onset, stage). GFAP and S-100B add little.

## INTRODUCTION

The translation of biomarkers from research validation to clinical implementation represents one of medicine’s most persistent challenges. Neurofilament light chain (NfL), a 68-kDa neuronal cytoskeletal protein released following axonal damage [7, 22], exemplifies this translational gap: extensively validated as a prognostic biomarker for ALS in research settings [1, 2, 3, 10, 12, 14] and recognized by the FDA as a surrogate endpoint, where NfL reduction supported the 2023 accelerated approval of tofersen [4, 23, 30] but the C9orf72 oligonucleotide BIIB078, which lowered neither neurofilament nor clinical decline, was discontinued [43]; yet it remains incompletely characterized in everyday clinical practice.

The foundational validation by Lu and colleagues in 2015 established blood NfL’s diagnostic and prognostic utility, demonstrating serum sensitivity of 89% and specificity of 75% for distinguishing ALS from healthy controls, with paired CSF correlation of r = 0.78 [1]. Gaiani et al. (2017) confirmed NfL’s diagnostic and prognostic value across ALS subtypes [2]. Verde et al. (2019) extended these findings using SIMOA technology to demonstrate serum NfL discrimination of ALS from disease controls [10], and the Khalil et al. (2018) review established NfL as a cross-disease biomarker of axonal injury [22]. Earlier SIMOA [10, 12, 14] and CSF studies [11, 15, 16, 18, 19], together with a cross-disease meta-analysis spanning ALS, frontotemporal dementia, and Alzheimer disease [20], established the underlying biology that the commercial platform now translates to routine use.

These validation studies, however, employed research conditions that bear little resemblance to clinical practice. The CReATe Consortium used SIMOA technology, achieved complete data capture through protocol-mandated assessments, and enrolled carefully selected patients [3]. Real-world implementation data remain limited; Davies et al. (2023) found serum NfL had limited diagnostic value in a real-world Oxford cohort, with nearly a quarter of cases showing serum NfL in the normal range at presentation (sensitivity 0.77) [5].

Critically, the commercially available Roche Elecsys ECLIA platform yields values 6-to 8-fold lower than SIMOA. Mondesert et al.’s 2025 four-platform comparison in ALS patients confirmed strong cross-platform agreement (R^2^ ≥ 0.939) and equivalent diagnostic AUCs (0.889–0.912) despite a 6-to 8-fold absolute difference [26], and Ashrafzadeh-Kian et al. (2024) reported similar between-assay differences in head-to-head plasma comparisons [27]. Both, however, are analytical comparisons that report elevated-NfL rates rather than assessments of clinical utility. Booth et al. (2025) generated age-specific reference intervals for the ECLIA assay [29].

Karam (Muscle & Nerve 2026) recently provided the first peer-reviewed clinical-utility assessment of ECLIA-based serum NfL, in peripheral neuropathy at Penn (n = 128) [28]. That study did not include ALS and used only NfL.

We therefore analyzed commercial ECLIA-based serum NfL as the primary marker, GFAP secondarily, and S-100B where data were available, in real-world ALS practice, in 58 chart-confirmed patients spanning the diagnostic and management continuum at a tertiary academic center. The primary objective was to test whether ECLIA NfL retains its correlation with the ALSFRS-R decline rate under routine clinical conditions; secondary objectives were to define its platform-specific distribution relative to research SIMOA assays and to evaluate whether serum GFAP or S-100B contributes diagnostic or prognostic information. We hypothesized that ECLIA NfL would track the progression rate despite real-world data fragmentation, whereas GFAP and S-100B would add little.

## METHODS

### Study Design

We conducted a retrospective analysis of real-world electronic health record data from a single tertiary academic center. The IRB-approved protocol included a waiver of informed consent for retrospective EHR review.

### Inclusion and Exclusion Criteria

Candidate patients were identified by ICD-10 coding (G12.21) combined with serum biomarker test ordering at Georgetown University Hospital between January 2022 and March 2026. Each candidate underwent individual chart review to confirm an ALS diagnosis. Patients whose records did not support ALS were reassigned to their actual diagnosis and excluded, leaving 58 chart-confirmed patients.

### Participant Characteristics

The cohort comprised adults under care for confirmed or suspected ALS, spanning the full diagnostic and management continuum from new presentations to established disease. Demographic and clinical characteristics are summarized in Table 1.

**Table 1.** ALS Cohort Demographics (n = 58)

| Characteristic | Value |
| --- | --- |
| Age, mean ± SD (years) | 63.7 ± 14.4 |
| Age, median (range) | 64 (27–92) |
| Sex, male | 30 (52%) |
| Sex, female | 23 (40%) |
| Sex, unknown | 5 (9%) |
| Race, African American (of those captured) | 17 of 53 (32%) |
| Race, White (of those captured) | 22 of 53 (42%) |
| Race, Other (of those captured) | 5 of 53 (9%) |
| Race, declined/unknown (of those captured) | 9 of 53 (17%) |
| Ethnicity, non-Hispanic (of those captured) | 35 of 53 (66%) |
| Ethnicity, Hispanic | 0 (0%) |
| Ethnicity, declined/unknown (of those captured) | 18 of 53 (34%) |

### Sampling Procedures

Patients were sampled from routine clinical practice, not a research protocol. Biomarkers were ordered at the discretion of the treating clinician, so per-modality availability varied across the cohort (Supplementary Table S1). We accepted the variable pre-analytical conditions inherent to clinical care rather than imposing research-grade collection protocols.

### Sample Size, Power, and Precision

No a priori power calculation was performed. The sample size was fixed by the number of eligible patients in the study period, all of whom were included. Subset sizes vary by analysis according to data availability (Supplementary Table S1), which limits the precision of the longitudinal and multi-biomarker estimates.

### Measures and Covariates

All biomarker measurements were performed by LabCorp (Burlington, NC) using the Roche Elecsys ECLIA platform. NfL was assayed via test code 505115 (analytical sensitivity 0.241 pg/mL, inter-assay CV <5%); GFAP and S-100B were measured on the same platform. Where the laboratory information system recorded multiple entries for a single draw on the same calendar date, these were treated as one timepoint; only measurements on different calendar dates were considered repeat draws for longitudinal analysis.

Item-level ALSFRS-R scores were extracted from structured clinical-note templates. For patients with assessments on two or more dates, the rate of decline was calculated as (first ALSFRS-R − last ALSFRS-R) divided by months of follow-up, in points lost per month. Patients were classified as fast progressors (>1 point/month decline), intermediate (0.5–1 point/month), or slow (<0.5 point/month). For the analysis of NfL against time from symptom onset, progression rate was also expressed as the ΔFS index, calculated as (48 − ALSFRS-R) divided by months from symptom onset [35], using the same cut-points. The serial decline rate (measured between assessments) underlies the NfL–decline-rate correlation; ΔFS (anchored at symptom onset) classifies progression rate in the time-from-onset trajectory.

Treatment and genetic information were ascertained from available clinical notes. Upper-versus lower-motor-neuron predominance was assigned from documented examination findings (upper: spasticity, hyperreflexia, extensor plantar responses; lower: atrophy, fasciculations, hyporeflexia). Age, disease stage, and time from symptom onset were recorded as covariates for the GFAP analysis.

### Statistical Analysis

Spearman rank correlation assessed the association between first-per-patient NfL and the ALSFRS-R decline rate, with a sensitivity analysis restricting NfL to within 90 days of the rate-defining assessments. Between-group comparisons used the Mann-Whitney U test. Within-subject NfL trajectories were summarized descriptively. Within-patient GFAP slopes were estimated for patients with two or more draws using a within-patient (fixed-effects) estimator pooled across patients. SIMOA-equivalent values were estimated using the published Elecsys-to-SIMOA regression of Booth et al. (SIMOA = 6.566 × Elecsys −0.812; r = 0.991) [29], consistent with the 6-to 8-fold difference reported by Mondesert et al. [26]. Cross-disease comparison used concurrent MS, CIDP, SMA, and ATTR amyloidosis cohorts measured on the identical ECLIA platform. P < 0.05 was considered significant.

Serum GFAP was examined against age, the ALSFRS-R decline rate, disease stage (48 minus the contemporaneous ALSFRS-R score), and time from symptom onset, using first-per-patient values. To separate disease-related change from normal aging, GFAP was age-adjusted to a reference age of 60 years using a published healthy-control rate of approximately 1.5% per year; an internal estimate gave a concordant result. The fraction of GFAP values within the non-ALS comparator range was determined against the 90th percentile of the peripheral-neuropathy and unclassified cohorts. For Figure 2, age-adjusted GFAP was plotted against months from symptom onset with serial draws connected within patients; the left panel is a conceptual illustration of accumulation at three fixed rates and is not derived from these data.

## RESULTS

### Cohort Characteristics

The final cohort comprised 58 patients with chart-confirmed ALS (Table 1). The median age was 64 years (range 27–92; mean 63.7 ± 14.4); 30 patients (52%) were men, 23 (40%) women, and 5 (9%) of unrecorded sex. Among the 50 patients with available clinical notes, disease-modifying treatment was documented as riluzole in 9, edaravone in 10, and AMX0035 (sodium phenylbutyrate–taurursodiol) in 2. A small number of patients with SOD1-associated ALS were receiving tofersen.

Consistent with real-world data, the available measurements differed by patient and analysis (Supplementary Table S1). Fifty-seven patients had at least one NfL value (80 total), 29 had at least one GFAP value (39 total), and 14 had at least one S-100B value (18 total). Forty-two patients had at least one ALSFRS-R assessment (190 total), of whom 32 had a calculable decline rate; 31 had both an NfL value and a decline rate, and 13 had NfL on two or more distinct dates. Individual NfL values for all patients and draws are listed in Supplementary Table S4.

### Serum NfL Distribution

First-per-patient serum NfL ranged from 0.96 to 62.20 pg/mL, with a median of 7.06 (IQR 4.06–17.30), a mean of 11.1, and a coefficient of variation of 99%, spanning the breadth of disease states encountered in practice (Table 2). Applying the published ECLIA-to-SIMOA conversion [29] places these values at a median of roughly 46 pg/mL (IQR ≈ 26–113).

**Table 2.** Serum NfL Distribution: First-per-Patient (n = 57)

| Statistic | ECLIA (LabCorp) | SIMOA-equivalent* |
| --- | --- | --- |
| Median (pg/mL) | 7.06 | ~46 |
| IQR (pg/mL) | 4.06–17.30 | ~26–113 |
| Min–Max (pg/mL) | 0.96–62.20 | ~5–408 |
| Mean (pg/mL) | 11.1 | ~72 |
| CV | 99% | — |

*\*SIMOA-equivalent via Booth et al. regression ( $SIMOA = 6.566 \times Elecsys - 0.812$ ; $r = 0.991$ ) [29]; Elecsys runs 6- to 8-fold lower than SIMOA [26].*

### NfL and ALSFRS-R Progression Rate

Among the 31 patients with both a first-per-patient NfL value and a calculable ALSFRS-R decline rate, NfL correlated with the rate of functional decline (Spearman r = 0.704, p < 0.0001; Figure 1). The association was essentially unchanged when NfL was restricted to within 90 days of the slope-defining assessments (n = 17, r = 0.809, p < 0.0001), indicating it was not an artifact of non-contemporaneous sampling. Stratified by progression rate, fast progressors (>1 ALSFRS-R point/month, n = 8) had a mean NfL of 17.10 pg/mL versus 4.64 pg/mL in slow progressors (<0.5 point/month, n = 21), a 3.7-fold difference (Supplementary Table S2). NfL did not differ by race (White median 6.62 versus African American 7.85 pg/mL, p = 0.94) or by upper-versus lower-motor-neuron predominance (5.8 versus 6.3 pg/mL, p = 0.79), favoring neither compartment, and was only weakly, non-significantly associated with age (Spearman r = 0.23, p = 0.10), arguing against demographic confounding. Adjusting for age and sex left the NfL–decline-rate association essentially unchanged (partial Spearman r = 0.73, p < 0.0001), and NfL was unrelated to static disease stage (48 − ALSFRS-R; r = 0.26, p = 0.13), confirming the relationship reflects the rate of disease activity rather than age or accumulated severity, in contrast to GFAP, which was strongly age-dependent.

**Figure 1.**
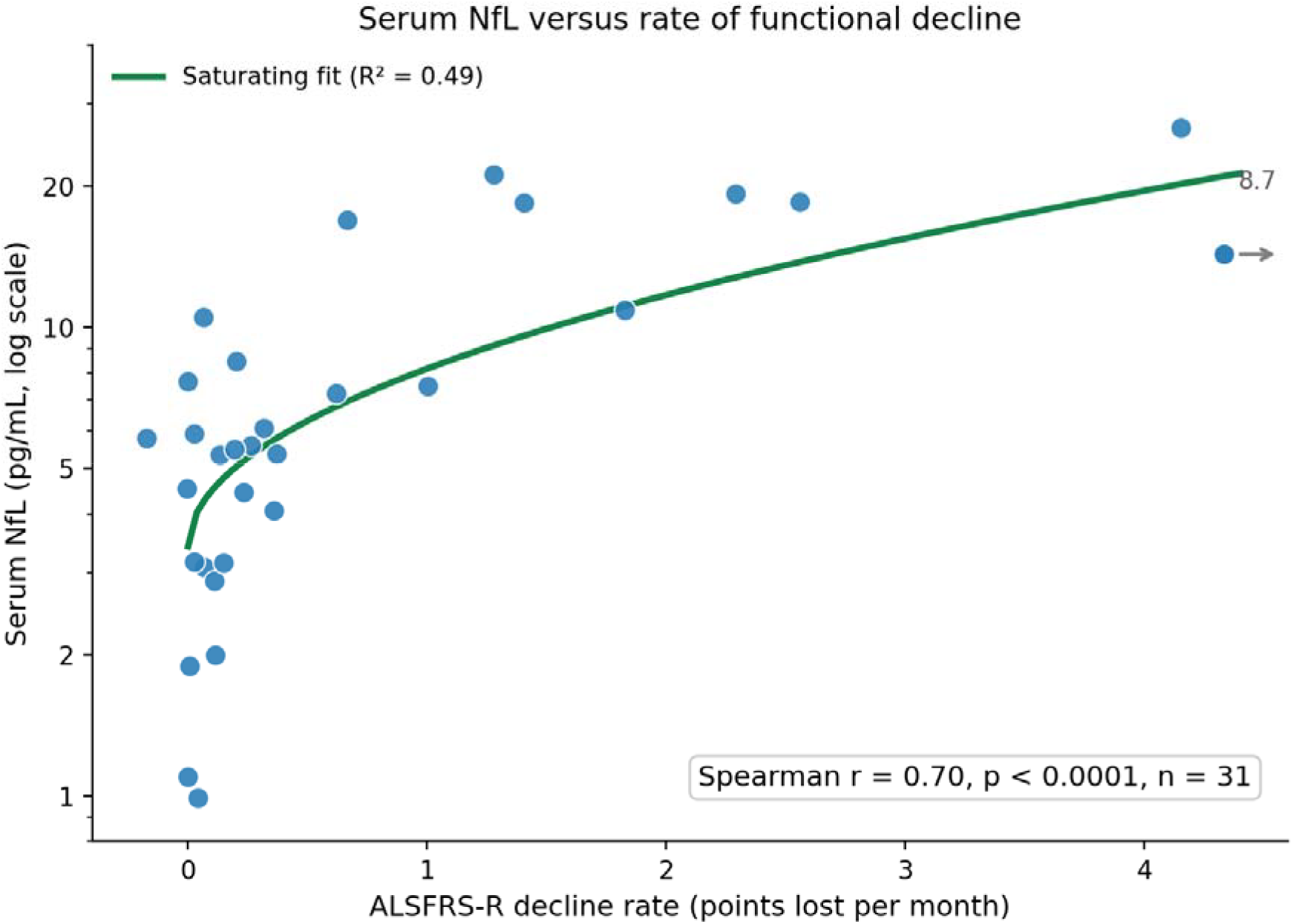
First-per-patient serum NfL versus the ALSFRS-R decline rate (points lost per month; zero indicates stable disease), n = 31; Spearman r = 0.704, p < 0.0001. The fitted curve is an illustrative saturating model: NfL rises steeply at low decline rates and plateaus. One patient with a decline rate of 8.7 points/month is off scale and marked at the right axis margin (arrow). Fast progressors (n = 8) and slow progressors (n = 21) are defined in Results.

### Longitudinal NfL

Thirteen patients had serial NfL on two or more distinct dates, with within-subject trajectories including both increases and decreases.

### GFAP and S-100B

GFAP (serum and plasma combined) was available in 29 patients (39 values; median 71.70 pg/mL) and S-100B in 14 patients (18 values; median 53.05 pg/mL) (Supplementary Table S3). Most GFAP values were not elevated: 24 of 29 (83%) fell within the range of the non-ALS neurologic comparators. GFAP correlated strongly with age (Spearman r = 0.77, p < 0.001) but not with the ALSFRS-R decline rate (r = −0.13, p = 0.65, n = 15), disease stage (r = 0.12, p = 0.61, n = 20), time from symptom onset (r = 0.09, p = 0.76, n = 13; Figure 2), or upper-versus lower-motor-neuron predominance (median 30.8 versus 68.3 pg/mL, p = 0.13).

**Figure 2.**
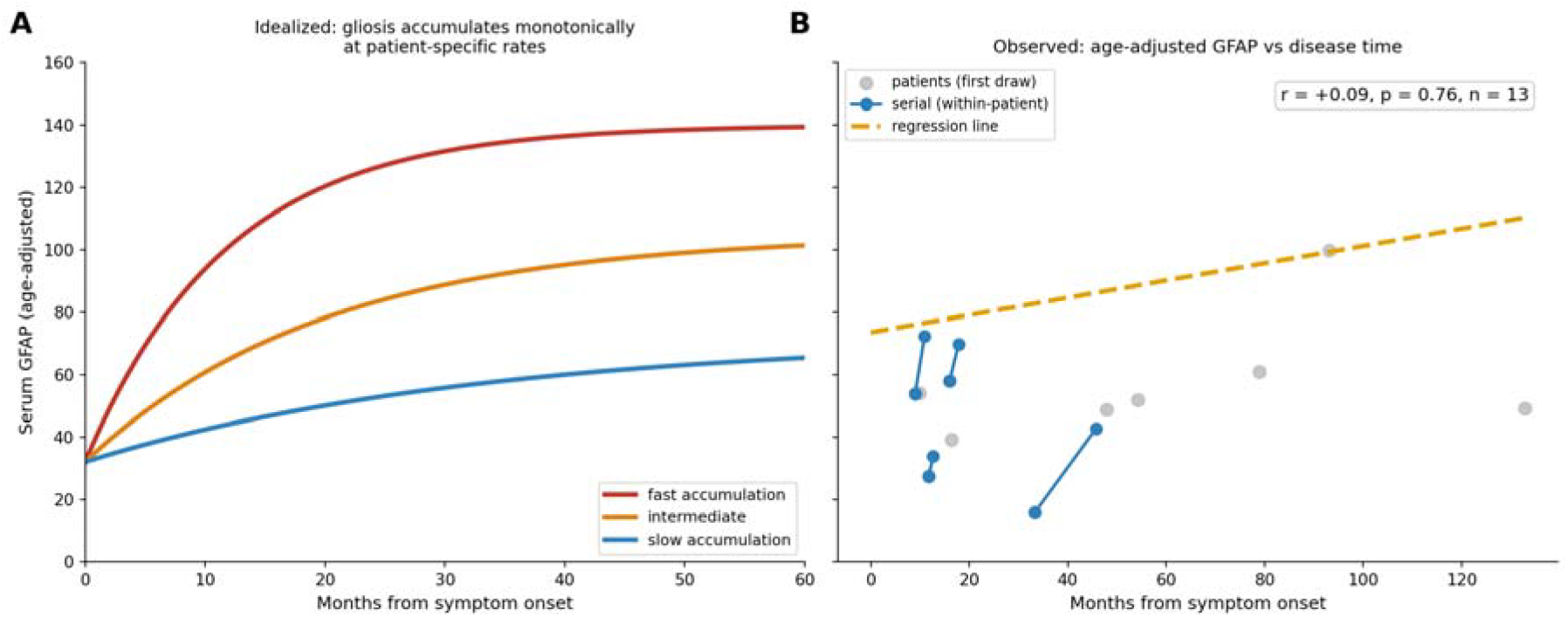
Serum GFAP and disease time. (A) Conceptual model: reactive astrogliosis accumulating from onset at fast, intermediate, and slow rates (illustration, not cohort data). (B) Observed age-adjusted GFAP (to age 60) versus months from symptom onset; grey points are first-per-patient values and blue segments join serial draws within a patient (regression not significant, r = +0.09, p = 0.76). Heterogeneous accumulation sampled at different disease times yields a flat cross-section despite the upward within-patient drift.

Among patients with two or more draws, GFAP rose or was stable in most and fell in none, but the within-patient change was small and not significant (pooled slope p = 0.11). Age-normalization left no association with disease stage (r = −0.11) or disease time (r = −0.08). Paired NfL–GFAP analysis was limited by the smaller number of patients with both markers.

## DISCUSSION

The principal finding is that serum NfL measured on the commercial ECLIA platform retains its prognostic relationship with ALS progression rate in routine clinical practice. Despite the heterogeneity, sparse longitudinal sampling, and incomplete documentation of real-world data, baseline ECLIA NfL correlated with the ALSFRS-R decline rate, and the association persisted when restricted to measurements drawn within 90 days of the defining assessments. The additional markers added little: serum GFAP scaled with age rather than the motor disease and showed no significant association with progression rate, stage, or motor-neuron predominance, a non-specific pattern, while S-100B showed no clear value in a preliminary subset.

This confirms, on the commercial platform, the prognostic framework established in research SIMOA cohorts, that baseline serum NfL predicts functional decline and survival [1, 3], even though a substantial minority of ALS patients present with NfL in the reference range [5]. To our knowledge this is among the first clinical-utility characterizations of ECLIA NfL in ALS; prior ECLIA data derive largely from analytical platform comparisons [26, 27].

### NfL as a Marker of Axonal Injury Rate

The inverse correlation between baseline NfL and subsequent decline reflects the marker’s biology. With a serum half-life near three weeks, NfL indexes the rate of axonal degeneration integrated over the preceding weeks, so a steady-state concentration predicts the future trajectory more faithfully than it tracks a contemporaneous ALSFRS-R score [32]. The serial data make the corollary concrete: the change in NfL over time is more informative than any single value; a rising concentration signals an accelerating injury rate, a stable low value indolent disease [17]. The marker is best read as a trajectory, not against a fixed cutoff.

A falling serum NfL in a deteriorating patient must not be read as improvement. Late in the course, progressive loss of the large motor axons that release NfL can lower the marker even as disability accumulates, decoupling it from clinical state. Plasma neurofilament heavy chain falls toward end-stage in fast-progressing ALS [33], and serum NfL is lower in long-duration disease [34], both consistent with this, and cautioning against reading a late value or downward trend without disease-stage context. Such a decline has not, to our knowledge, been characterized for serum NfL in routine ECLIA practice.

### Source of the Serum Neurofilament Signal

Whether serum NfL reports a central or peripheral source bears on interpretation. One deeply phenotyped study read it as corticospinal because it rose with transcranial-magnetic-stimulation-defined upper-motor-neuron burden [36], yet NfL was lowest in primary lateral sclerosis, the most purely corticospinal phenotype. The intraspinal corticospinal tract is a thin neurofilament reservoir, so a central process may yield little NfL regardless of tempo. A peripheral source fits better: NfL derives mainly from large peripheral motor axons, reporting anterior-horn and peripheral degeneration, co-varying with motor-neuron involvement, as shown for cerebrospinal fluid neurofilaments [13], but lower in primary lateral sclerosis [25].

Phenotype-to-compartment mapping is leaky both ways: a lower-motor presentation may hide subclinical corticospinal involvement, and a non-evocable cortical response scored upper-motor-neuron can itself arise from anterior-horn depletion. Pure lower-motor labels are unreliable; corticospinal degeneration appears at autopsy in roughly half of clinically pure cases [39], and quantitative susceptibility mapping detects motor-cortex change across motor neuron disease [40], and in these records lower-motor signs were charted less than upper. Because serum markers register only substantial axonal loss, focal or early involvement stays subthreshold. NfL therefore indexes the rate of motor-axon degeneration wherever it occurs, not its compartment.

### Comparison with the Benatar Trajectory Model

Benatar and colleagues defined the temporal trajectory of NfL across the ALS continuum: in genetically at-risk individuals it rises months to years before the first weakness, continues through early symptomatic disease, and plateaus by the time established disease is diagnosed, remaining relatively stable within an individual thereafter [8, 32]. The plateau reflects the rate of ongoing axonal injury and is strongly influenced by genotype, which also governs the timing of the initial rise [32]. Our cross-sectional correlation with progression rate fits this framework: a higher steady-state level marks a faster injury rate [9].

Benatar’s trajectories rise and plateau but do not descend, yet in practice we observed both stable and clearly declining serial values, not a contradiction but a consequence of the setting. Disease-modifying therapy lowers the injury rate: tofersen reduces NfL in SOD1-ALS [4, 23, 30], so a treated level can fall rather than plateau. Diagnostic heterogeneity adds mimics whose biomarker profile need not match ALS (see Clinical Implications). And opportunistic sampling, unlike protocol-driven measurement, spans many patients across genotypes, rates, stages, and treatments rather than one within-patient arc.

### Platform-Specific Interpretation

The roughly 6-to 8-fold difference between commercial ECLIA and research SIMOA values has direct diagnostic weight: applying SIMOA-derived thresholds [3, 6] to an ECLIA result risks misclassifying a rapid progressor as lower-risk. ECLIA-specific interpretation is required, with age-specific reference intervals [29] as a starting framework. Even in ALS, the highest-NfL motor neuron disease on this platform [21, 22], many values fall within or near the reference range; prognostic information lies in relative position and longitudinal change, not an elevated-versus-normal split.

### Multi-Biomarker Profiling: GFAP and S-100B

S-100B showed no discriminative value and is confounded by non-neural sources. GFAP marks reactive astrogliosis [41] but clears from blood within days, rising acutely after traumatic brain injury [42], so it registers only after acute, massive injury, not the slow diffuse astrogliosis of ALS. Accordingly, cohort GFAP scaled strongly with age, not the motor disease: most values were normal, and neither rate, stage, duration, nor predominance predicted it, consistent with the amyloid co-pathology that raises GFAP in older adults, though amyloid was not measured here [37, 38].

GFAP was not purely static: within patients it rose, in one several-fold faster than the age-expected rate, while a presymptomatic patient with floor-level NfL stayed flat, a small activity-dependent component too sparse to quantify (Figure 2). The same age-driven decoupling appears on LUMIPULSE [36], so convergence across three platforms argues biology, not artifact. Serum may also blunt or lag an astroglial signal that CSF captures more faithfully, given slow release and blood–brain-barrier gating [41]. Serial NfL remains the actionable ALS readout; S-100B adds nothing, whereas a markedly elevated GFAP may still flag concurrent age- or amyloid-related astroglial co-pathology.

### Clinical Implications

A low NfL during an ALS workup argues against an active, rapidly degenerative process [5] and should prompt reconsideration of the differential, though it identifies no single alternative. Because NfL indexes axonal injury, a low value is equally compatible with an indolent motor disorder, a primary muscle disease, or a presymptomatic gene carrier [8, 24], and inflammatory neuropathies can themselves run normal-to-high [28]. NfL therefore reconsiders the diagnosis rather than confirming or excluding any specific mimic, and is most informative alongside electrodiagnostic, antibody, and genetic testing.

For monitoring, the trajectory matters more than any single value [32]: a rising serial NfL signals an accelerating injury rate or, in a treated patient, inadequate disease control [31]. A low value should not be read as quiescence, since few remaining axons in advanced or slowly progressive disease can yield a low NfL despite substantial deficits [34]. Because commercial ECLIA values run well below SIMOA [26, 27], monitoring should use a consistent platform and ECLIA-specific, age-adjusted reference intervals rather than thresholds derived elsewhere.

### FUTURE DIRECTIONS

Read against the right question, serum NfL works on three scales: a near-binary diagnostic flag in early disease, a presymptomatic trajectory in at-risk carriers, and a continuous prognostic rate marker. Diagnostically, an elevated value supports active motor-neuron degeneration and a normal one argues against it, helping separate ALS from indolent mimics, though imperfect sensitivity means a normal result cannot exclude the diagnosis [5]. The profile is also non-monotonic, low before symptom onset and rising thereafter, so it must be read against disease timing. In at-risk gene carriers, serial NfL can flag rising axonal pathology before phenoconversion [8, 24]. For progression, serial NfL can track the trajectory on or off therapy, though drug-response monitoring is unproven; because NfL reports the rate of ongoing axonal loss, late or burnout disease may read low despite severe deficits.

Integration is the next step: serum NfL combined with complementary biomarkers and brain imaging, in particular quantitative susceptibility mapping for the upper-motor-neuron component [40], alongside digital biomarkers and structured clinical metrics, built into multimodal and ultimately federated multi-center models that differentiate ALS from its mimics and support both diagnosis and individualized prognosis.

### Limitations

This single-center retrospective analysis draws different questions from different patient subsets (Supplementary Table S1). Genetic and treatment data came from clinical notes and likely under-represent true status, and disease duration at the NfL draw was often unextractable. Fewer than a third of patients had a repeat NfL draw, since routine care does not reliably re-test, the main constraint on the trajectory analyses. Renal function and body mass also influence NfL [6], but documented renal disease was rare and recorded body mass largely normal, so neither likely affected the results.

Electrodiagnostic documentation was incomplete, so the upper-versus lower-motor-neuron assignment is coarse and the Penn Upper Motor Neuron Score could not be reconstructed. The within-patient GFAP analysis rests on six patients and was not significant. We did not measure amyloid, phosphorylated tau, TDP-43 species, or a peripheral panel, markers linked to the GFAP signal and the lower-motor-neuron compartment [36, 37, 38], so the anatomical source of serum GFAP cannot be localized here.

## CONCLUSIONS

Commercial ECLIA brings neurofilament light chain into routine ALS care: the prognostic relationship established on research SIMOA assays survives real-world fragmentation, so the marker is usable where patients are managed rather than only in research cohorts. Its clinical meaning lies in the trajectory, not the absolute value: a rising NfL marks active progression, a persistently low one slow or indolent disease, read against an ECLIA-specific reference range rather than SIMOA-derived thresholds.

Serum GFAP and S-100B add little: S-100B is uninformative, and GFAP tracks age and amyloid co-pathology [37, 38] more than the motor disease. Because NfL is shaped by genotype, genetic testing remains worthwhile even when no variant is identified. The marker’s fullest value will come combined with advanced motor-cortex neuroimaging and emerging biomarkers.

## Ethics approval

This retrospective study was approved by the Georgetown University Institutional Review Board (protocol no. STUDY00008842; initially approved October 10, 2025), which granted a waiver of informed consent for review of existing electronic health record data.

## Data availability

De-identified data supporting the findings are available from the corresponding author on reasonable request, subject to institutional data-sharing agreements.

## Use of AI

The author used an AI assistant (Anthropic Claude) for copy-editing and manuscript preparation, and reviewed and verified all content, taking full responsibility for the work.

## Author contributions

N.S.S. is the sole author and is responsible for all aspects of the work, including conception, data acquisition and analysis, drafting, and final approval.

